# Machine Learning Based Prediction of COVID-19 Mortality Suggests Repositioning of Anticancer Drug for Treating Severe Cases

**DOI:** 10.1101/2021.11.11.21266048

**Authors:** Thomas Linden, Frank Hanses, Daniel Domingo-Fernández, Lauren Nicole DeLong, Alpha Tom Kodamullil, Jochen Schneider, Maria J.G.T. Vehreschild, Julia Lanznaster, Maria Madeleine Ruethrich, Stefan Borgmann, Martin Hower, Kai Wille, Thorsten Feldt, Siegbert Rieg, Bernd Hertenstein, Christoph Wyen, Christoph Roemmele, Jörg Janne Vehreschild, Carolin E. M. Jakob, Melanie Stecher, Maria Kuzikov, Andrea Zaliani, Holger Fröhlich, LEOSS study group

**Affiliations:** Fraunhofer Institute for Algorithms and Scientific Computing (SCAI), Schloss Birlinghoven, 53757 Sankt Augustin, Germany; University of Bonn, Bonn-Aachen International Center for IT, Friedrich Hirzebruch-Allee 6, 53115 Bonn, Germany; Emergency Department, University Hospital Regensburg, 93053 Regensburg, Germany; Clinic and Polyclinic for Internal Medicine II, Technical University of Munich, 81675 Munich, Germany; Department II of Internal Medicine, Infectious Diseases, University Hospital Frankfurt, Goethe University, 60590 Frankfurt, Germany; Department of Internal Medicine II, Hospital Passau, Innstraße 76, 94032 Passau, Germany; Institute for Infection Medicine and Hospital Hygiene, University Hospital Jena, 07743 Jena, Germany; Department of Infectious Diseases and Infection Control, Hospital Ingolstadt, 85049 Ingolstadt, Germany; Department of Pneumology, Infectious Diseases and Intensive Care, Klinikum Dortmund gGmbH, Hospital of University Witten / Herdecke, 44137 Dortmund, Germany; University Clinic for Haematology, Oncology, Haemostaseology and Palliative Care, Johannes Wesling Medical Centre Minden, 32429 Minden, Germany; Department of Gastroenterology, Hepatology and Infectious Diseases, University Hospital Düsseldorf, Medical Faculty of Heinrich Heine University Düsseldorf, Moorenstrasse 5, 40225 Düsseldorf, Germany; Department of Medicine II, University Hospital Freiburg, 79110 Freiburg, Germany; Christoph Wyen, Praxis am Ebertplatz Cologne, 50668 Cologne, Germany; Internal Medicine III - Gastroenterology and Infectious Diseases, University Hospital Augsburg, 86156 Augsburg, Germany; Department I for Internal Medicine, University Hospital of Cologne, University of Cologne, 50931 Cologne, Germany; Fraunhofer Institute for Translational Medicine and Pharmacologie (ITMP), VolksparkLabs, Schnackenburgallee 114, 22535 Hamburg, Germany; Department for Infectious Diseases and Infection Control, University Hospital Regensburg, Germany

**Keywords:** precision medicine, covid19, machine learning, drug repositioning, explainable ai

## Abstract

Despite available vaccinations COVID-19 case numbers around the world are still growing, and effective medications against severe cases are lacking. In this work, we developed a machine learning model which predicts mortality for COVID-19 patients using data from the multi-center ‘Lean European Open Survey on SARS-CoV-2-infected patients’ (LEOSS) observational study (>100 active sites in Europe, primarily in Germany), resulting into an AUC of almost 80%. We showed that molecular mechanisms related to dementia, one of the relevant predictors in our model, intersect with those associated to COVID-19. Most notably, among these molecules was tyrosine kinase 2 (TYK2), a protein that has been patented as drug target in Alzheimer’s Disease but also genetically associated with severe COVID-19 outcomes. We experimentally verified that anti-cancer drugs Sorafenib and Regorafenib showed a clear anti-cytopathic effect in Caco2 and VERO-E6 cells and can thus be regarded as potential treatments against COVID-19. Altogether, our work demonstrates that interpretation of machine learning based risk models can point towards drug targets and new treatment options, which are strongly needed for COVID-19.

## 1. Introduction

As of October 2021, the ongoing SARS-COV-2 pandemic led to almost 5 million reported deaths worldwide according to data from the for US Institute for Health Metrics and Evaluation (https://covid19.healthdata.org/). In addition, economic costs are estimated to reach the order of several trillion dollars for the USA alone (Cutler and Summers 2020). While effective vaccinations are now available, there are still a considerable number of infected people worldwide. Moreover, effective medications for treating severe cases are still scarce. Remdesivir, a drug originally developed against the Ebola virus, is currently the only approved COVID-19 drug in the European Union, and evidence suggests that it has little effect on the overall survival of COVID-19 patients (Ansems et al. 2021).

Several studies have revealed general risk factors for a poor disease outcome, such as age, male gender, and low platelet count (Gaze 2020; O’Driscoll et al. 2021; Qeadan et al. 2021; Wool and Miller 2021). In addition, machine learning (ML) models have been published to predict mortality risk for individual patients, primarily based on data from Intensive Care Units and electronic health records from the US and UK (Ali et al. 2021; Banoei et al. 2021; Gao et al. 2020; Jones et al. 2021; Ryan et al. 2020; Schwab et al. 2021; Vaid et al. 2020) as well as a few other countries (Kar et al. 2021; Mahdavi et al. 2021). Notably the 4C mortality score developed by Ali et al. based on data from the UK has recently been validated within an intendent study in Canada (Jones et al. 2021). None of these models have resulted in a change of clinical routine or the identification of new treatment options so far.

In this work, we specifically investigated data from nearly 5,700 PCR or rapid test confirmed SARS-COV-2 patients recruited in more than 100 European active sites, primarily all over Germany. For these patients, disease symptoms, vital parameters, biomarkers from urine and blood, and diagnosed comorbidities were available. Using these data and ML, we first developed a model that can predict mortality with an area under receiver operator characteristic curve (AUC) of almost 80% up to 60 days in advance. One of the relevant predictors in our model was a prior diagnosis of dementia, which increases the mortality risk by about 15%. Based on this finding, we explored the overlap between COVID-19, Alzheimer’s (AD), and Parkinson’s Disease (PD) molecular disease mechanisms, which pointed us to tyrosine kinase 2 (TYK2) as a potential new drug target. Finally, our experimental data with Caco2 and VERO-E6 cells suggests that Sorafenib and Regorafenib, two approved anti-cancer drugs, could be repositioned for treating severe COVID-19 cases.

## 2. Results

### 2.1. Overview about LEOSS Data

The Lean European Open Survey on SARS-CoV-2 infected patients (LEOSS - https://leoss.net/) is an observational, multi-center study focusing on PCR or rapid test confirmed patients. Study centers are primarily University Medical Centers, but also include other hospitals, institutes, and medical practices. Active sites cover several European countries but have a primary focus on Germany. They are thought to generate representative data of (primarily hospitalized) COVID-19 cases, at least for Germany. In order to ensure anonymity in all steps of the analysis process, an individual LEOSS Scientific Use File (SUF) was created, which is based on the LEOSS Public Use File (PUF) principles described in (Jakob et al. 2020). The baseline data from more than 100 active sites, collected at time of a positive test or diagnosis, comprises patient demographics, disease symptoms, vital parameters, biomarkers from urine and blood, and comorbidities. Follow-up information, including survival, was available for patients between 18 and 85 years. These data were further filtered to only include patients with less than 50% of missing data at baseline, resulting into n = 5,679 patients (Table 1). Out of those 5,679 patients, 5225 (92%) were inpatient, and 569 (10.0%) were reported death cases within a follow-up period of up to 78 days. Among them, 430 (76% of 569) patients were reported death cases within the first 20 days (Figure 1).

**Table 1:**
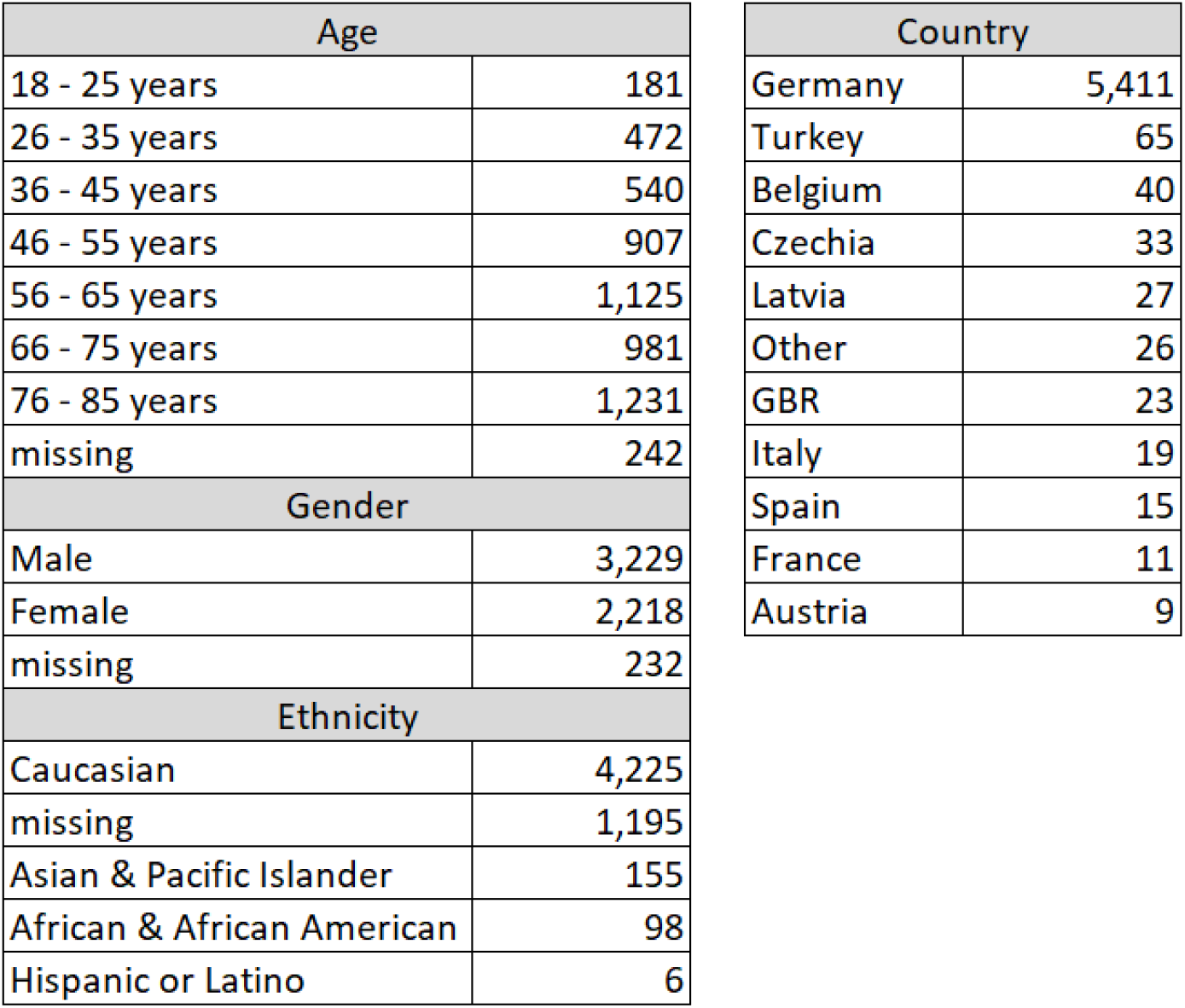
Overview of patient demographics in LEOSS.

| Age |  | Country |  |
| --- | --- | --- | --- |
| 18 - 25 years | 181 | Germany | 5,411 |
| 26 - 35 years | 472 | Turkey | 65 |
| 36 - 45 years | 540 | Belgium | 40 |
| 46 - 55 years | 907 | Czechia | 33 |
| 56 - 65 years | 1,125 | Latvia | 27 |
| 66 - 75 years | 981 | Other | 26 |
| 76 - 85 years | 1,231 | GBR | 23 |
| missing | 242 | Italy | 19 |
| Gender |  | Spain | 15 |
| Male | 3,229 | France | 11 |
| Female | 2,218 | Austria | 9 |
| missing | 232 |  |  |
| Ethnicity |  |  |  |
| Caucasian | 4,225 |  |  |
| missing | 1,195 |  |  |
| Asian & Pacific Islander | 155 |  |  |
| African & African American | 98 |  |  |
| Hispanic or Latino | 6 |  |  |

**Figure 1:**
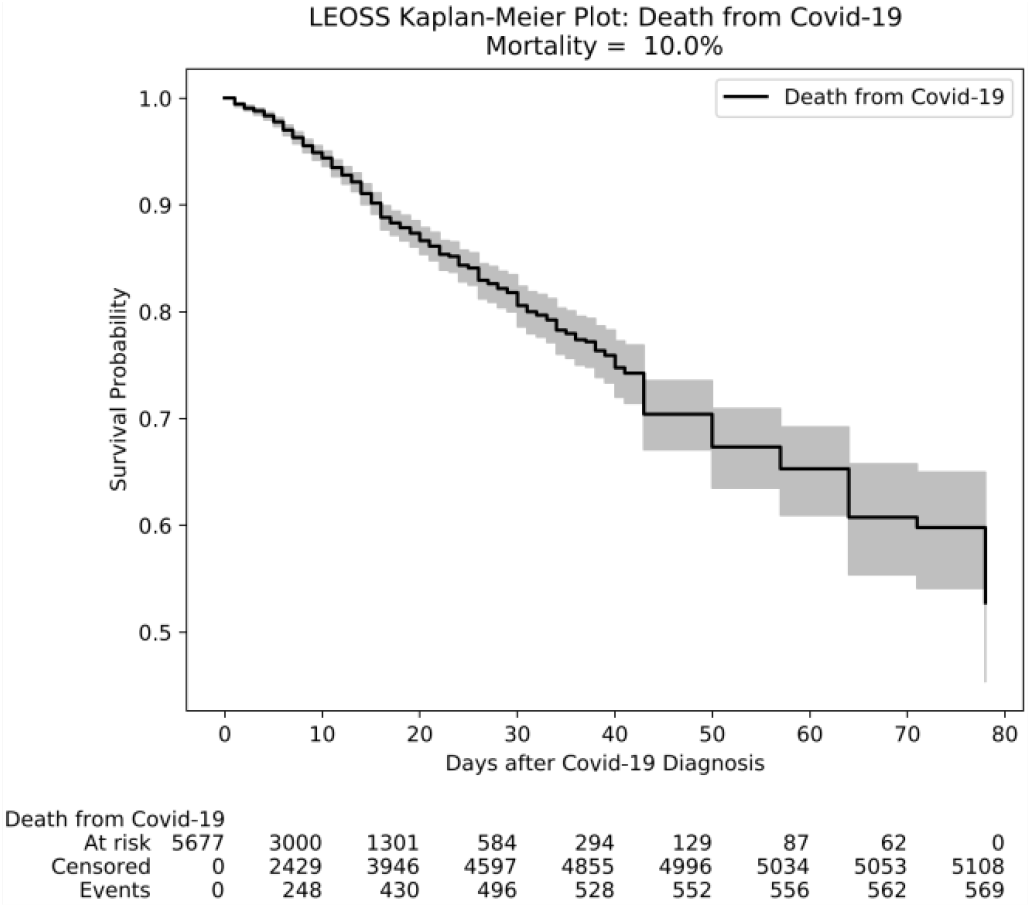
Kaplan-Meier plot of COVID-19 patients in LEOSS.

### 2.2. Machine Learning Can Predict Mortality with High Accuracy

We implemented and compared a broad panel of time-to-event machine learning models to predict patient survival using only LEOSS baseline data:

- Elastic net penalized Cox proportional hazards regression (Cox 1972; Zou and Hastie 2005; Wu 2012)
- Elastic net penalized Weibull accelerated failure time regression (Weibull 1951; Zou and Hastie 2005; Khan and Shaw 2013)
- DeepSurv – a neural network approach using a loss function derived from a Cox proportional hazard model (Katzman et al. 2018)
- Random Survival Forests (Ishwaran et al. 2008)
- XGBoost Survival Embeddings – a popular stochastic gradient boosting algorithm using a loss function derived from a Weibull regression (Vieira et al. 2021)

We evaluated models via a five-fold cross-validation (CV). In other words, we split the entire dataset into five outer folds, and we subsequently left out one of these folds for testing the model, while the rest of the data was used for model training and tuning. Notably, splitting of the data was performed in a stratified manner, such that the number of events was equally maintained across all folds. We tuned the hyper-parameters within the CV loop using an extra level of inner five-fold CV (see Section 4.1 for details). We employed Uno’s C-index as a metric to assess prediction performance (Uno et al. 2011). A C-index of 50% indicates chance level, whereas a C-index of 100% would reflect a perfect concordance of model predictions and observed death cases in the test data (see Section 4.2 for details).

Overall, elastic net penalized Weibull regression achieved the best discrimination performance with ∼77% C-index and low calibration error (Integrated Brier Score – IBS) of 0.12 (Figure 2, **Supplementary Table 1**). Therefore, this algorithm was used to subsequently train a final model on the entire dataset while using the previously described approach for hyper-parameter tuning.

**Figure 2:**
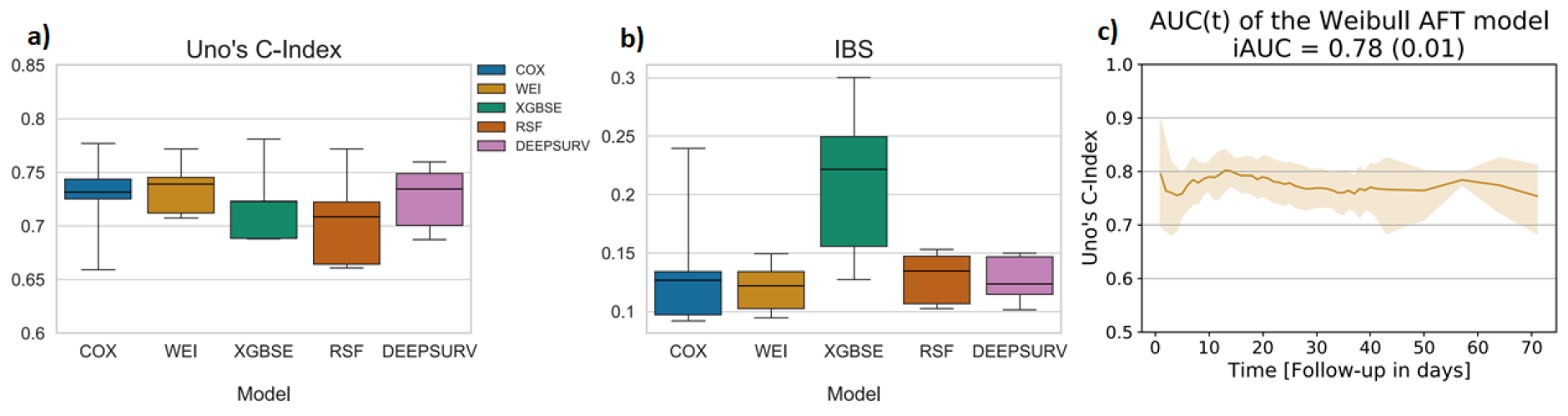
(a) Model prediction performance measured via Uno’s C-index on held out test sets (COX = elastic net penalized Cox proportional hazards regression; WEI = elastic net penalized Weibull accelerated failure time regression; XGBSE = XGBoost Survival Embeddings; RSF = Random Survival Forest; DEEPSURV = DeepSurv); (b) model calibration error measured via Integrated Brier Score (IBS) on held out test sets; (c) model prediction performance as function of time on held out test sets with 95% confidence interval, with integrated AUC (iAUC) denoting the mean (standard error) AUC over time.

### 2.3. Diagnosis of Dementia as a Relevant Predictor

The final model was further explored with respect to the impact of most relevant predictors using Shapley Additive Explanations - SHAP (Lundberg and Lee 2017). Briefly, SHAP is an approach from cooperative game-theory to decompose the overall prediction of the model into a sum of individual feature contributions (see details in 4.3). In total, the final model comprised 160 features. A complete list can be found in **Supplementary File 1**.

Figure 3a shows the most influential features according to SHAP, while Figure 3b summarizes the influence of entire feature modalities, indicating that lab measures were the most relevant type of features (23.5% cumulative importance). Disease symptoms ranked second (20.5%) and comorbidities third (13.2% cumulative importance). Age, gender, platelet count as well as elevated troponin and ferritin concentrations were among the top predictors in the model, which are all known risk factors (Gaze 2020; O’Driscoll et al. 2021; Qeadan et al. 2021; Wool and Miller 2021). Comorbidity associated predictors included hypertension, an acute kidney injury, diabetes and dementia (**Supplementary Table 2, Supplementary File 2**). Again, this is concordant with the current literature (Corona et al. 2021; Du et al. 2021; Paek et al. 2020).

**Figure 3:**
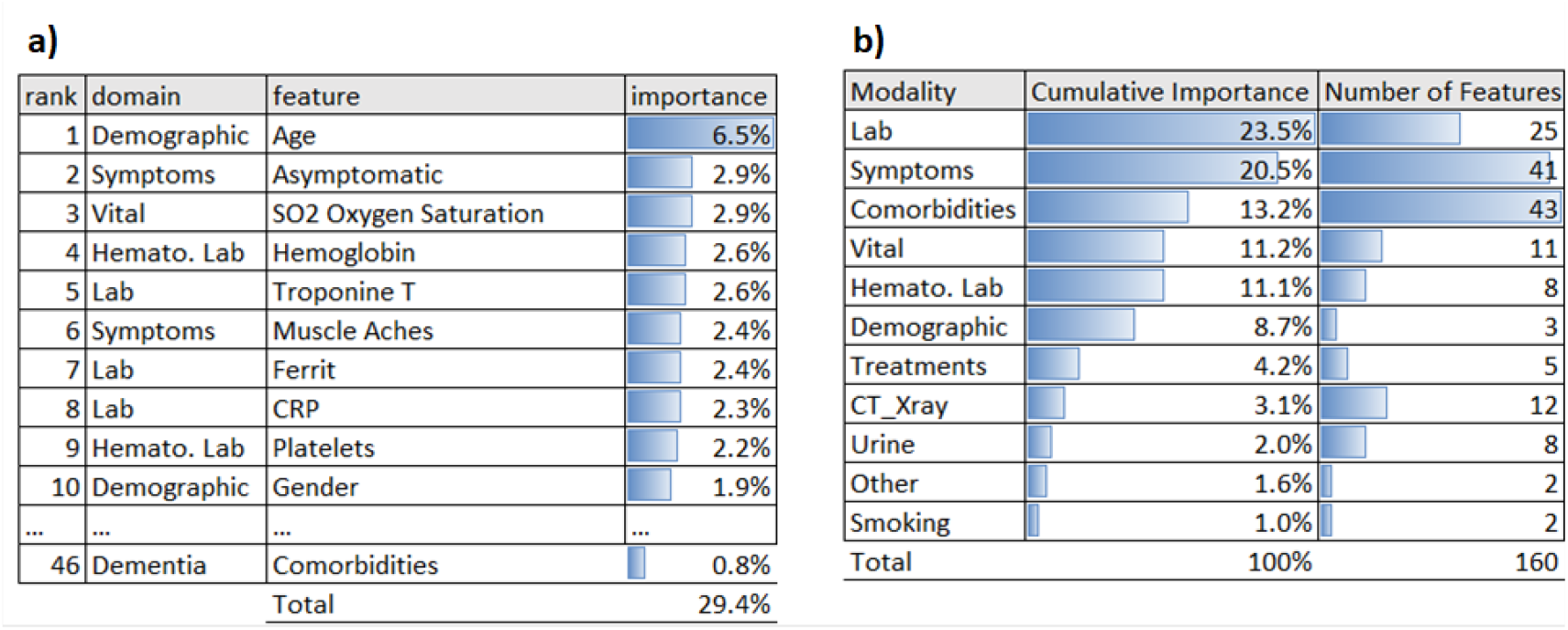
Feature importance using absolute SHAP values: (a) top 10 predictors; (b) cumulative influence per feature modality

Figure 4 displays partial dependency plots for several relevant predictors, describing the relationship between individual feature attributes and their impact on estimated hazard ratios. Accordingly, a prior diagnosis of dementia results into an ∼15% increased mortality risk after COVID-19 infection (hazard ratio dementia vs. non-dementia: ∼1.15; 95% CI: [1.08, 1.24]). Notably, there are different possible explanations for this finding: (a) dementia might be a proxy for age; (b) dementia might, independently of age, trigger biological, physiological and psychological mechanisms that contribute to an unfavorable disease outcome.

**Figure 4:**
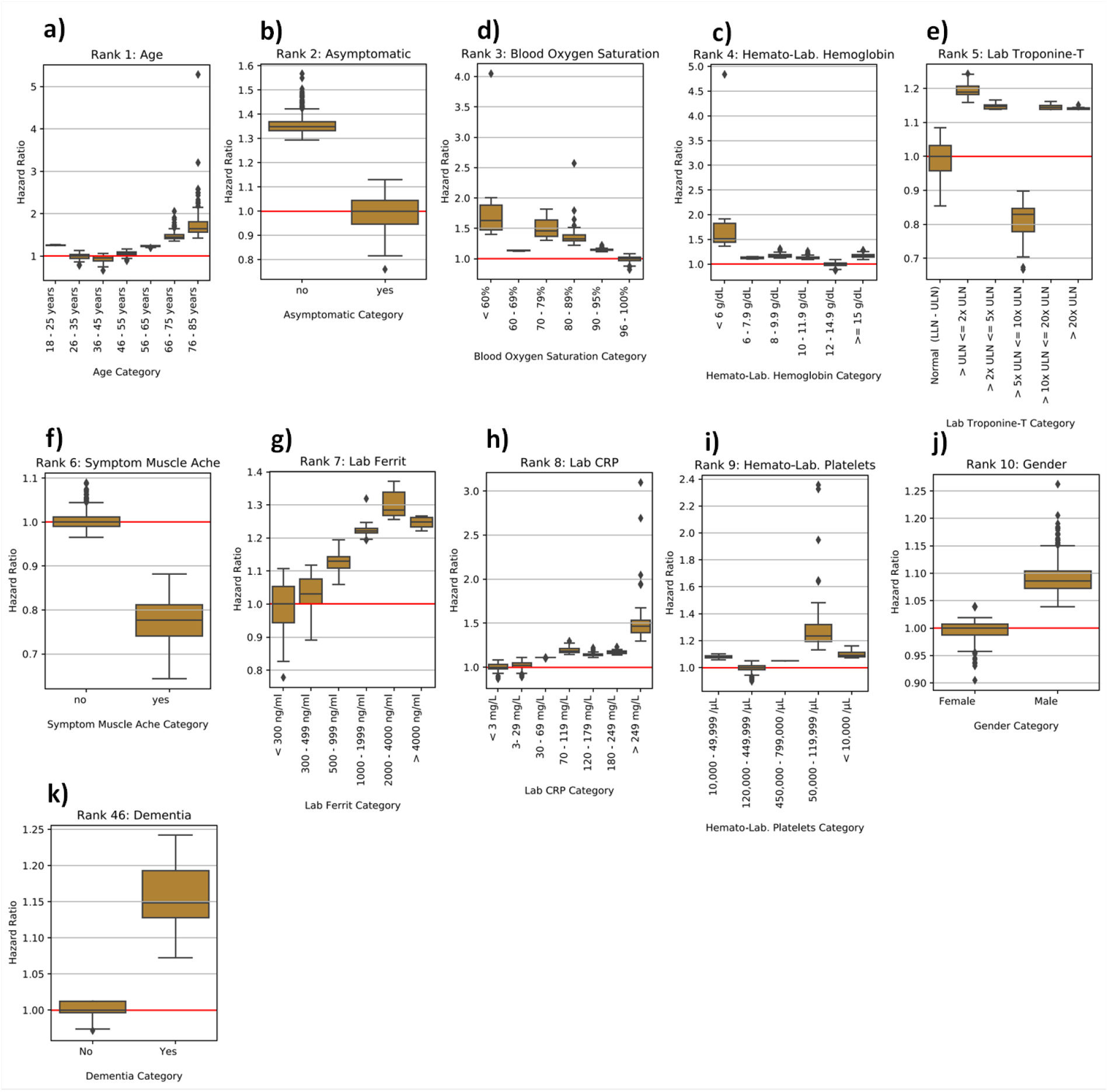
Partial dependence plots for most influential predictors. Boxplots show the distribution of patient specific hazard ratios per variable category. The red horizontal line defines the reference. The hazard ratio describes by which factor the median lifetime is expected to change compared to reference.

### 2.4. Commonly Affected Molecular Mechanisms Between Neurodegenerative Disorders and COVID-19

We aimed for a more in-depth exploration of potential overlaps of neurodegeneration and COVID-19 disease mechanisms. Notably, there has been increasing evidence that SARS-COV-2 can enter the central nervous system (Boldrini, Canoll, and Klein 2021; Krasemann et al. 2021; Meinhardt et al. 2021), raising the question of potential interactions with dementia disease pathologies. In this context, (Zhou et al. 2021) recently reported an overlap of transcriptionally dysregulated biological pathways in a very limited number of patients with Alzheimer’s Disease (AD) and COVID-19.

Here, we focused more broadly on shared molecular mechanisms linking COVID-19 with AD as well as Parkinson’s Disease (PD), another major neurodegenerative disorder, which has previously been associated with an increased risk for an unfavorable outcome of a SARS-COV-2 infection (Scherbaum et al. 2021; Vignatelli et al. 2021). By looking at the intersection between AD and PD cause-and-effect models (referred as knowledge graphs -KGs) and the corresponding COVID-19 KG, in this work we found a series of mechanisms that were shared between all three disease etiologies **(Supplementary Table 3)**.

Firstly, one of the mechanisms identified by our approach is related to three proteins involved in the innate immune system (i.e., DDX58, MAVS, and IFIH1), and more specifically in the detection and response to viruses. These proteins are involved in both indications. For example, MAVS interacts with the RNA helicase RIG-I/MDA-5 after the dsRNA of the virus is recognized, leading to the initiation of the antiviral signaling cascade (Zheng et al. 2020). Related with this process is the second shared mechanism, which corresponds to the activation of the inflammasome and the subsequent triggering of caspase activation through cytokine secretion. This mechanism has been strongly linked with both AD (Hansen, Hanson, and Sheng 2018) and PD (Wang et al. 2019) as well as COVID-19 (Pan et al. 2021). In the context of neurodegeneration, the activation of the inflammasome leads to the secretion of inflammatory cytokines and cell death through pyroptosis, to which both AD and PD are associated via tangle and plaque formation and death of dopamine neurons, respectively (Anderson et al. 2021). Similarly, in the context of COVID-19, the inflammasome is activated by the proteins of the SARS-CoV-2 virus, which in turn leads to the production of inflammatory molecules, and in some cases leads to hyperinflammation (Pan et al. 2021). Finally, TYK2 is also present in all three KGs. It is known to be implicated in the regulation of apoptosis in the amyloid cascade of AD (Wan et al. 2010) as well α-synuclein-induced neuroinflammation and dopaminergic neurodegeneration (Qin et al. 2016).

Lastly, IL-6 and IL-10 are among two of the interleukins secreted after inflammasome activation, one of the shared mechanisms between these pathologies, and their increased expression has been shown to be predictive of COVID-19 severity (Dhar et al. 2021). Furthermore, the interaction between two other proteins (i.e., DDIT3 and BCL2L11) involved in the regulation of apoptosis is also suggested as a common mechanism across these indications (Fricker et al. 2018; Inde et al. 2021).

### 2.5. Sorafenib and Regorafenib as Potential Treatments Against COVID-19

In the following, we specifically focused on TYK2, which is a protein involved into the amyloid cascade. TYK2 inhibition results into effective regulation of IFNα, IL-10, IL-12, and IL-23 (He et al. 2019), which has specifically been reported in neurodegenerative disorders (Porro, Cianciulli, and Panaro 2020). TYK2 has been patented as drug target in AD (CN102112879B, China, (Ip et al. 2015)). In addition, genetic variants in TYK2 have recently been associated to COVID-19 disease severity (COVID-19 Host Genetics Initiative 2021). Moreover, we found several kinase inhibitors active against SARS-Cov2 in a cellular screen for anti-cytopathic effect (anti-CPE) in two different cellular environments: Caco2 (Ellinger et al. 2021) and VERO-E6 (Zaliani et al. 2020).

We challenged VERO-E6 cells with SARS-CoV2 pretreated with compounds from the Fraunhofer Repurposing Library (5632 compounds - https://www.itmp.fraunhofer.de/en/innovation-areas/drug_screening_repurposing.html), the EUOS Bioactives library (∼2500 compounds - https://ecbd.eu/compound/#lib{value='2'}lib{value='2'}), and a proprietary “Safe in Man” library of compounds having passed phase I clinical trial (∼600 compounds). In VERO-E6 cells, only Regorafenib showed a clear antiviral CPE potency with an IC50 of around 3 – 5 uM. In Caco2 cells, Sorafenib and Regorafenib demonstrated a similar antiviral CPE potency with an IC50 of around 1uM for both molecules (Figure 5).

**Figure 5.**
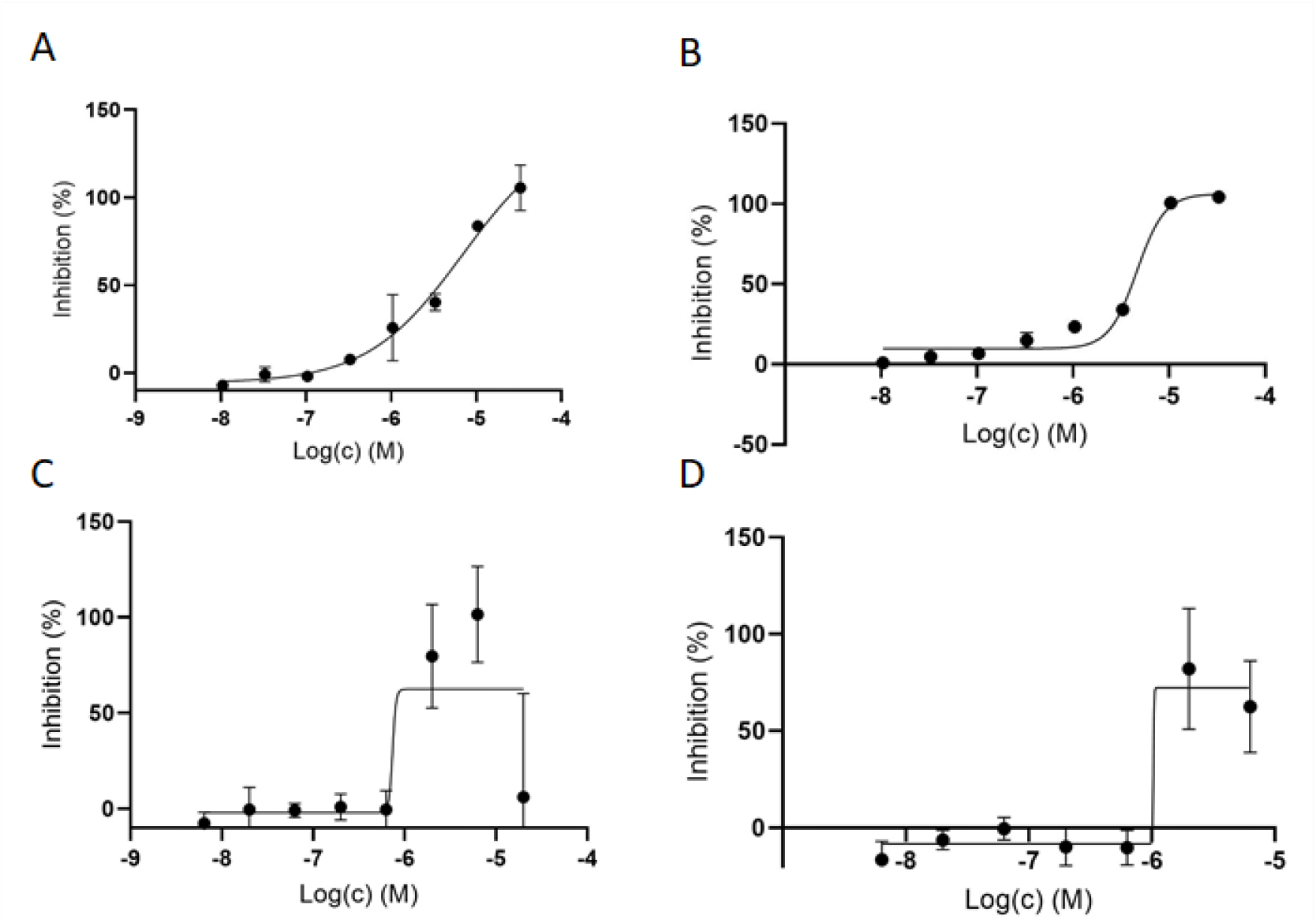
Regorafenib (panels A and C) and Sorafenib (panels B and D) activities measured in different cell lines (Vero-E6 cells upper panels; Caco2 cells lower panels) as percentage inhibition of viral cytopathic effect normalized to Remdesivir as positive control (100%). Cells in wells have been treated with SARS CoV-2 virus and drugs and, after 72 hours after infection, were stained, washed and counted if alive.

## 3. Conclusion

As of October 2021, the rates of completely vaccinated individuals in many Western countries are stagnating between 60 – 70%, while the fraction of vaccinated individuals is globally only around 36% (Mathieu et al. 2021). Correspondingly, case numbers in many countries around the world are still increasing. Hence, there is an unmet need for effective and cost-efficient medications against severe cases.

In this work, we first developed a highly predictive ML model for predicting COVID-19 mortality on an individual patient basis using deep observational data from LEOSS, primarily covering the inpatient situation in Germany (95% of patients). To our knowledge, this is the first ML based mortality model based on such (notionally) representative German data. Notably, ML models predicting alternative endpoints using LEOSS have been published recently (Jakob et al. 2021; Werfel et al. 2021).

Our ML model demonstrates similar prediction performance to the well-known 4C mortality score, which has been developed based on representative data from the UK (Ali et al. 2021). However, a direct comparison between both models is not possible, because the 4C model is formulated as a classifier predicting all-cause in-hospital mortality, whereas our model is formulated as a time-to-event model predicting all-cause time dependent mortality risk after COVID-19 diagnosis. Our model, thus, considers censoring of survival times after patients have left hospital or other medical facilities. Our mortality model was built on a set of patients, which is thought to be primarily representative for German hospitals. Whether there are unknown selection biases, remains an open question and they were not under our control. Moreover, it is unclear whether our model would be predictive for patients in other countries.

We showed that dementia, as one of the relevant predictors in our model, intersects on a molecular mechanism level with COVID-19. Together with evidence from recent GWAS studies, this pointed us to TYK2 as a potential drug target for COVID-19. Using a cellular screening assay for anti-cytopathic effect, we identified the anti-cancer drugs Regorafenib and Sorafenib as potential drug candidates against COVID-19. Notably, the known association of JAK family inhibitors like Regorafenib and Sorafenib with cellular inflammatory cytokines can be further characterized by investigating transcription dynamics within the first 12 hours after SARS-COV-2 viral infection compared to mock control (Stukalov et al. 2021). Based on such data, Stukalov et al. tested both compounds in the A549-ACE2 cell line and reported increased virus growth after treatment. Other authors recently reported Sorafenib to be a potent STING inhibitor effectively stopping virus growth in THP1 cells and thus suggested to pay more attention to COVID-19 treatment strategies that address the dysregulation of cytokines (Deng, Yu, and Pei 2020). Since the used cell lines in both cases were different from ours, results are not directly comparable. Hence, we see a need for further tests with Regorafenib and Sorafenib in other cell systems.

In addition to further experimental validation of Regorafenib and Sorafenib, it could be interesting to explore in large scale clinical real-world data whether SARS-COV-2 infected patients treated with Regorafenib or Sorafenib demonstrate a lower mortality than other SARS-COV-2 patients.

Overall, our work demonstrates that interpretation of an ML based risk model trained on rich data can point towards drug targets and new treatment options, which are strongly needed for COVID-19.

## 4. Methods

### 4.1. Machine Learning Models for Predicting COVID-19 Mortality

We compared five different machine learning algorithms, as outlined in Section 2.2. Here, we only elaborate on the best performing one, namely the elastic net penalized Weibull regression: The elastic net is a regularization and variable selection method, to shrink coefficients using a linear combination of *L*_1_ and *L*_2_ penalties. The Weibull regression is an accelerated failure time (AFT) model, which means that covariates act multiplicatively on (survival) time. It is used when the proportional hazards assumptions from the Cox model are not satisfied and/or to directly estimate (the effect of covariates on) expected failure times, where the time until failure is the duration of survival.

Let *i* = {1, …,*n*}denoting the *i*-th patient, model from the AFT family define a log-linear relationship between failure time *T*_*i*_ and a patients p-dimensional covariate vector *x*_*i*_, coefficients *β* and i.i.d. errors *ε*_*i*_ ∼*F*, where *F* is a statistical distribution of choice:

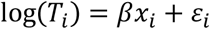

This can also be expressed as 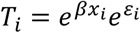. Which means that, if 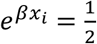 then aging is accellerated to as twice as normal, and if 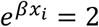 then aging is only half as normal. Depending on the choice for *F*, a particular model type from this family is specified, e.g. a Weibull AFT model.

Parameters of a standard Weibull AFT model can be estimated via maximum likelihood (Zhang 2016). In our case coefficients β are additionally penalized via the elastic net penalty:

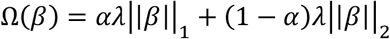

Hyperparameters (i.e. α, λ) were tuned with Bayesian hyperparameter optimization using the Optuna package (Akiba et al. 2019) within the inner-loop of the nested cross-validation. Early stopping was used if applicable (DeepSurv, GBM, XGBSE), and the best candidate model was subsequently selected. We chose the 5-fold cross-validated Harrell’s C-index (Harrell et al. 1982) as objective for the hyperparameter tuning. We ran the optimization for twenty initial epochs, adopted the search space if reasonable, and then ran it for another twenty rounds. Thus, forty hyperparameter sets were evaluated and the resulting best combination was selected based on the highest objective function value. Using this hyperparameter set, we subsequently trained a model on the entire training data and evaluated it on the held-out test set.

### 4.2. Uno’s Concordance-Index

The prediction performance of time-to-event models can be evaluated with respect to discriminating between subjects with different event times via Uno’s C-index (Uno et al. 2011): The C-index (Concordance index) is a generalization of the area under receiver operator characteristic curve (AUC) for time-to-event models (Heagerty and Zheng 2005; Schmid and Potapov 2012). A value of 100% means perfect discriminative performance, and 50% is comparable to random predictions.

In essence, Uno’s C is a rank correlation between the risk predictions and the observed event times. The C-index measures the concordance across all pairs of patients (*i,j*), *i* ≠ *j*. A pair is classified concordant if the predicted risk is higher for the patient with lower survival time. Uno’s C-index was developed as an alternative to Harrell’s C-index in settings with high censoring rates and leads to consistent concordance estimates under the general random censoring assumption. Uno’s C-index uses an inverse probability censoring weighting (IPCW) approach (Uno et al. 2011):

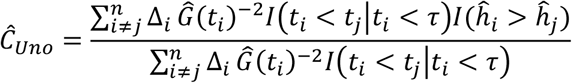

The numerator counts the concordant pairs and the denominator the valid pairs, respectively. For patients *i* = {1, …,*n*}, Δ_*i*_ is 1 if an event (death) was observed and otherwise 0, 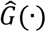 is the Kaplan-Meier estimator for the *censoring distribution* for IPCW, 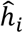 is the risk prediction of the *i* -th patient, *t*_*i*_ is the observed time and τ is a stability parameter, for further details see (Uno et al. 2011).

### 4.3. Feature Importance using SHAP

Shapley Additive Explanations (Lundberg and Lee 2017) are a model-agnostic approach from coalitional game theory. The assumption of this framework is, that individuals (feature attributes) are cooperating as a team (patient feature vector) for a joint outcome (model prediction). SHAP’s goal is to estimate those individual contributions to the outcome. Key properties are a) the solution is unique; b) local exactness, which means the sum of feature contributions matches the output; c) if a feature has no impact, then it’s SHAP-value is zero.

Mathematically, additivity and property b) can be described as:

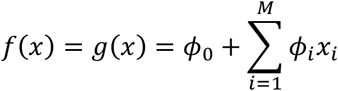

with *f*(*x*) being the original model, g(*x*) the explanation model and *ϕ*_*i*_ the SHAP value of the *i* -th feature for the model input vector *x* and *ϕ*_*0*_ denoting the expectation value of *f*(*x*). In other words: The SHAP values *ϕ*_*i*_ quantifies how much a particular feature pushes the prediction away from the population average *ϕ*_*0*_. SHAP values *ϕ*_*i*_ are computed as follows:

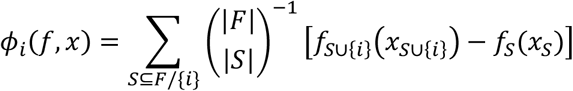

In other words, SHAP values are defined as a weighted (binomial coefficient) sum of the differences between (in square brackets) “prediction including the feature” minus “prediction excluding the feature”, for any subset in the power set *F*.

### 4.4. Confidence Intervals for Hazard Ratios

To construct a confidence interval for the hazard ratio of “dementia vs. non-dementia” we performed a bootstrap: We resampled 100,000 times with replacement a pair of a demented and non-demented patient. We then calculated the ratio of the SHAP values for the feature “prior dementia diagnosis” for both patients.

### 4.5. Identification of Common Molecular Mechanisms Between COVID-19 and neurodegenerative diseases

To identify the shared molecular mechanisms between COVID-19, AD, and PD, we leveraged several resources listed in **Supplementary Table 3**. These were combined into two independent Knowledge Graphs (KGs) following the harmonization procedure described in our previous work (Schultz et al. 2021) and (Domingo-Fernández et al. 2017). By doing so, we combined disease specific molecular interactions pertaining to COVID-19 and two neurological indications (i.e., AD and PD) into graph structures: one for COVID-19 and one for AD and PD. Subsequently, we calculated the intersection of these graphs. **Supplementary File 2** contains the corresponding shared mechanisms as an Excel table.

## Data Availability

The data analyzed in this study is subject to the following licenses/restrictions: Data will be available for research purposes. Data access has to be requested from https://leoss.net/ Access can be requested from https://leoss.net/registration/

https://github.com/thomasmooon/leoss-cov19

https://leoss.net/

## Code availability

The source code of the analyses presented in this paper is available at https://github.com/thomasmooon/leoss-cov19.

## Acknowledgments

We express our deep gratitude to all study teams supporting the LEOSS study. The LEOSS study group contributed at least 5 per mille to the analyses of this study: University Hospital Regensburg (Frank Hanses), Technical University of Munich (Christoph Spinner), University Hospital Frankfurt (Maria Vehreschild), Hospital Passau (Julia Lanznaster), University Hospital Jena (Maria Madeleine Ruethrich), Hospital Ingolstadt (Stefan Borgmann), Klinikum Dortmund gGmbH (Martin Hower), Johannes Wesling Hospital Minden Ruhr University Bochum (Kai Wille), University Hospital Duesseldorf (Bjoern-Erik Jensen), University Hospital Freiburg (Siegbert Rieg), Hospital Bremen-Center (Bernd Hertenstein), Practice at Ebertplatz Cologne (Christoph Wyen), University Hospital Augsburg (Christoph Roemmele), University Hospital Schleswig-Holstein Luebeck (Jan Rupp), Robert-Bosch-Hospital Stuttgart (Katja Rothfuss), Catholic Hospital Bochum (St. Josef Hospital) Ruhr University Bochum (Kerstin Hellwig), Elisabeth Hospital Essen (Ingo Voigt), Hospital Maria Hilf GmbH Moenchengladbach (Juergen vom Dahl), Marien Hospital Herne Ruhr University Bochum (Timm Westhoff), Municipal Hospital Karlsruhe (Christian Degenhardt), University Hospital Heidelberg (Uta Merle), University Hospital Munich/ LMU (Michael von Bergwelt-Baildon), Hospital Leverkusen (Lukas Eberwein), University Hospital Wuerzburg (Nora Isberner), University Hospital Essen (Sebastian Dolff), Hospital St. Joseph-Stift Dresden (Lorenz Walter), Hospital Ernst von Bergmann (Lukas Tometten), University Hospital Erlangen (Richard Strauss), University Hospital Cologne (Norma Jung), University Hospital Tuebingen (Siri Göpel), Bundeswehr Hospital Koblenz (Dominic Rauschning), Hacettepe University (Murat Akova), University Hospital of Giessen and Marburg (Janina Trauth), Hospital Fulda (Philipp Markart), National MS Center Melsbroek (Marie D’Hooghe), Evangelisches Hospital Saarbruecken (Mark Neufang), Malteser Hospital St. Franziskus Flensburg (Milena Milovanovic), University Hospital Schleswig-Holstein Kiel (Anette Friedrichs), University Hospital Ulm (Beate Gruener), Center for Infectiology Prenzlauer Berg Berlin (Stephan Grunwald), Elbland Hospital Riesa (Joerg Schubert), University Hospital Saarland (Robert Bals), Clinic Munich (Wolfgang Guggemos), Agaplesion Diakonie Hospital Rotenburg (David Heigener), Hospital Braunschweig (Jan Kielstein).

The LEOSS study infrastructure group: Jörg Janne Vehreschild (Goethe University Frankfurt), Susana M. Nunes de Miranda (University Hospital of Cologne), Carolin E. M. Jakob (University Hospital of Cologne), Melanie Stecher (University Hospital of Cologne), Lisa Pilgram (Goethe University Frankfurt), Nick Schulze (University Hospital of Cologne), Sandra Fuhrmann (University Hospital of Cologne), Max Schons (University Hospital of Cologne), Annika Claßen (University Hospital of Cologne), Bernd Franke (University Hospital of Cologne) und Fabian Praßer (Charité, Universitätsmedizin Berlin).

## Author Contributions

Drafted the manuscript: HF, TL, DDF, AZ; initiated and guided the project: HF; implemented machine learning models: TL; computational drug target identification: DDF, LDL, ATK; experimental validation: MK, AZ. Other authors: acquisition and preparation of LEOSS data

## Conflict of Interest

The authors declare no competing interests.

## Ethics Approval

LEOSS is registered at the German Clinical Trials Register (DRSK, S00021145) and was approved by the leading Ethics Commitee No. 20-600 “Ethikkommission des Fachbereichs Humanmedizin der Johann-Wolfgang-Goethe-Universität Frankfurt am Main, 60590 Frankfurt, Germany”. For the anonymization procedure see (Jakob et al. 2020).

## Funding

This work has been funded via the ‘COPERIMOplus’ initiative and supported by the Fraunhofer ‘Internal Programs’ under Grant No. Anti-Corona 840266.

The LEOSS registry was supported by the German Centre for Infection Research (DZIF) and the Willy Robert Pitzer Foundation.

## Notes

### Competing Interest Statement

The authors have declared no competing interest.

### Author Declarations

LEOSS is registered at the German Clinical Trials Register (DRSK, S00021145) and was approved by the leading Ethics Commitee No. 20-600 "Ethikkommission des Fachbereichs Humanmedizin der Johann-Wolfgang-Goethe-Universitaet Frankfurt am Main, 60590 Frankfurt, Germany". For the anonymization procedure see (Jakob et al. 2020).

