## Supplemental for "Machine Learning Based Prediction of COVID-19 Mortality Suggests Repositioning of Anticancer Drug for Treating Severe Cases"

| | **Uno's C** | **IBS** | **D-Calibration Max** | **D-Calibration p-value** |
|---|---|---|---|---|
| **COX** | 0.75 (0.02) | 0.14 (0.01) | 0.10 (0.00) | 0.80 (0.09) |
| **DEEPSURV** | 0.75 (0.01) | 0.13 (0.00) | 0.09 (0.00) | 0.82 (0.08) |
| **RSF** | 0.71 (0.02) | 0.13 (0.00) | 0.10 (0.00) | 0.81 (0.08) |
| **WEI** | **0.77 (0.01)** | **0.12 (0.00)** | 0.10 (0.00) | 1.00 (0.00) |
| **XGBSE** | 0.74 (0.02) | 0.21 (0.01) | 0.09 (0.00) | 0.71 (0.09) |

**Supplementary Table 1:** Model prediction performance (mean and standard error) measured via Uno's C-index on held out test sets (COX = elastic net penalized Cox proportional hazards regression; WEI = elastic net penalized Weibull accelerated failure time regression; XGBSE = XGBoost Survival Embeddings; RSF = Random Survival Forest; DEEPSURV = DeepSurv). Model calibration error measured via Integrated Brier Score (IBS) on held out test sets and D-Calibration (Haider et al. 2018). D-Calibration Max is the maximum percentage deviation from the expected value. D-Calibration p-value from chi-square test checking for D-Calibration, if larger than 0.05 then the model is D-Calibrated.

| Comorbidity | Importance |
|---|---|
| Hypertension | 1.9% |
| Acute Kidney Injury | 1.3% |
| Diabetes (with end organ damage) | 0.8% |
| Other Sensitization | 0.8% |
| Dementia | 0.8% |
| Diabetes (without end organ damage) | 0.8% |
| Coronary Artery Disease | 0.7% |
| Myocardial Infarction | 0.6% |
| AV Block | 0.6% |
| Chron. Pulm. Disease | 0.6% |

**Supplementary Table 2:** SHAP feature importance of the top-10 comorbidities. The complete list with SHAP values for all features is contained in **Supplementary File 1**.

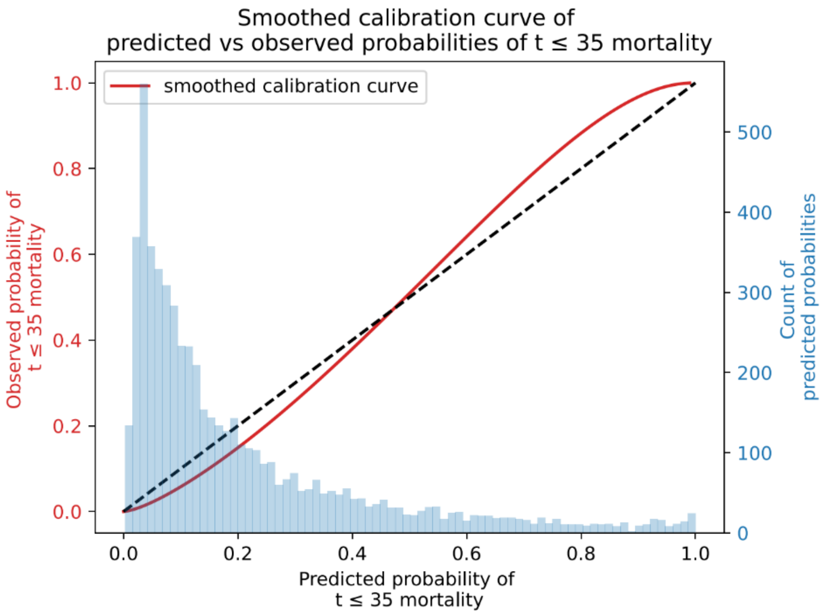

**Supplementary Figure 1:** Calibration of the final Weibull-AFT model trained. The difference between predicted probabilities (dashed black line) and observed probabilities (red curve) is less than around 10%.

| Resource Name | Reference | URL |
|---|---|---|
| NeuroMMSig Alzheimer's disease (AD) | PMID:28651363 | https://neurommsig.scai.fraunhofer.de/ |
| NeuroMMSig Parkinson's disease (PD) | PMID:28651363 | https://neurommsig.scai.fraunhofer.de/ |
| KEGG Alzheimer's disease pathway | PMID:10592173 | https://www.genome.jp/pathway/hsa05010 |
| KEGG Parkinson's disease pathway | PMID:10592173 | https://www.genome.jp/kegg-bin/show_pathway?hsa05012 |
| COVID-19 Knowledge Graph | PMID:32976572 | https://github.com/covid19kg and https://bikmi.covid19-knowledgespace.de |
| IntAct | PMID:33206959 | https://www.ebi.ac.uk/intact/query/annot:%22dataset:coronavirus%22?conversationContext=3 |
| BioGRID | PMID:30476227 | https://thebiogrid.org/project/3/covid-19-coronavirus.html |
| KEGG Coronavirus disease - COVID-19 | PMID:10592173 | https://www.genome.jp/pathway/hsa05171 |
| QIAGEN Corona Virus Network Explorer | PMID:33941085 | https://digitalinsights.qiagen.com/coronavirus-network-explorer/ |
| WikiPathways COVID-19 | PMID:32371892 | https://www.wikipathways.org/index.php/Portal:COVID-19 |

**Supplementary Table 3: List of resources employed to identify common molecular mechanisms between COVID-19 and neurodegenerative disorders.** The first column corresponds to the name of the resource. The second column includes the references of these resources. Finally, the third column points to the URLs where the resources are available. The table is divided between resources specific to neurodegenerative conditions (upper half) and COVID-19 (bottom half).

**Supplementary File 1: Excel including SHAP for all features in the final model.**

**Supplementary File 2: Excel including all common relations between the two KGs.**
